# Severe liver damage, low bone mineral content, and PDXDC1/TTC39B gene variations linked to NAFLD with metabolic syndrome

**DOI:** 10.1101/2023.05.11.23289665

**Authors:** Ankita Chatterjee, Bandana Mondal, Kausik Das, Dipankar Mondal, Ranajoy Ghosh, Abhijit Chowdhury, Priyadarshi Basu

**Author notes:** authors contributed equally (Ankita Chatterjee, Bandana Mondal). Corresponding author: Priyadarshi Basu.

## Abstract

**Background:** Fatty liver disease is a pathologic condition of liver due to excess deposition of fat in the liver and is considered as hepatic manifestation of metabolic syndrome (MetS). Metabolic syndrome is comorbid with multiorgan abnormalities and diseases. However, the phenotypic uniqueness and genetic background of fatty liver patients in the presence/absence of metabolic syndrome are unclear.

**Methods:** A cohort of 551 individuals with (FLD+) or without fatty liver (FLD-) was recruited.We subdivided the study cohort based on the presence or absence of metabolic syndrome (MetS) into four groups-Group4_FLD+/MetS+_; n=221, Group3_FLD+/MetS-_; n=39, Group2_FLD-/MetS+_; n=175 and Group1_FLD-/MetS-_ n=116. Pathophysiology was compared between the study groups. Association of genomic variants with Group4_FLD+/MetS+_; n=167 was studied compared to Group1_FLD-/MetS-_; n=74. The effects of the associated variants on gene expression were studied using eQTL mapping.

**Results and conclusions:** Among 551 individuals, 47.2% had fatty liver (FLD+) and 71.87% had metabolic syndrome (MetS+). Compared to Group3FLD+/MetS-, Group4FLD+/MetS+ patients had significantly higher age, higher adiposity, severe diabetic and lipid profile, liver damage marker, CRP, low bone mineral content and higher liver damage, both among the obese and the non-obese. Non-obese Group2FLD-/MetS+ patients had significantly higher serum TG and lower HDL, while obese Group3FLD+/MetS-patients had higher liver damage markers. Additionally, we also showed that in our population, Group4FLD+/MetS+ patients carried higher risk allele frequency in rs3761472-G(SAMM50,OR=2.9(2.0-4.1); p=0.002), rs738409-G(PNPLA3,OR=2.8(1.9-4.07)p=0.003),rs58542926-A(TM6SF2,OR=2.7(1.9-3.9)p=0.021),rs35665085-A(CECR5,OR=2.7(1.9-3.9)p=0.038),rs471364-G(TTC39B,OR=3.1(2.1-4.5)p=0.001),rs2800-G(SLC9A9,OR=3.1(2-4.5)p=0.028),rs7200543-A(PDXDC1,OR=2.261(1.1-4.8)p=0.031). Group4 patients with rs7200543-AA showed poor skeletal health. Thus, fatty liver with metabolic syndrome showed the most severe disease phenotype.

## Introduction

Worldwide, Fatty liver disease (FLD) in absence of viral infection affects approximately 25% of the total human population [1], [2] FLD is associated with liver failure, cardiovascular disease, chronic kidney disease, polycystic ovarian syndrome (PCOS) and hepatocellular carcinoma [3]. FLD can be classified as alcoholic liver disease (ALD) and non-alcoholic fatty liver disease (NAFLD; mild to moderate amount of alcohol consumption). Urbanization and sedentary lifestyle led to the gradual increase in the prevalence of NAFLD over the years [1] NAFLD is often associated with metabolic derangements [4], [5]. Metabolic syndrome (MetS), characterized by markers of obesity, hypertension, lipid profile, glycaemic profile and inflammatory markers, is the key risk factor associated with development and progression of FLD [5] [6]. The pathophysiology of NAFLD in the presence or absence of metabolic syndrome is highly variable, approximately 20%-25% of South Asians are affected with [7]. One study particularly mentioned that Chinese males were affected more with NAFLD coupled with metabolic syndrome compared to females [6], [8]. Hence, it is important to appropriately stratify the clinical features related to metabolic syndrome in NAFLD, to understand the disease pathophysiology and to design clinical trials [8].

A major component of metabolic syndrome is obesity [9], [10]. However, ours and others previous studies have shown that among Asian population, fatty liver disease is prevalent among non-obese population [10]–[13]. [13]. – although the disease severity among obese and non-obese patients were comparable [11], [14]. Approximately, 12.1% of the fatty liver disease were observed among the non-obese individuals and 39% of non-obese patients are generally affected with NASH [15]. It is unclear how metabolic syndrome is associated with fatty liver development, in absence of markers of obesity. Only two recent studies [16], [17] showed the prevalence of metabolic syndromes and associated fatty liver among the lean individuals. There is a serious gap in knowledge on the pathophysiology of fatty liver among the obese compared to the non-obese individuals.

Finally, like other complex disease traits, susceptibility to metabolic disorders is often related to genetic predisposition. Previous studies have shown that multiple variants were associated with traits like serum triglycerides, LDL, HDL, blood pressure, C-reactive protein, HbA1c and FBG [5]. Abnormal levels of each of these traits can contribute to the development of metabolic disorder [5]. Limited research was done to delineate the genetic underpinning of FLD in the presence/absence of MetS [8]. In our previous study, we have identified both common and unique SNPs associated hepatic fat content in the Indian population [18] differential association with the SNPs among the lean and non-lean population [18]. In this study, we took candidate gene approach to map the SNPs associated with fatty liver development among individuals with metabolic syndromes.

## Methods

### Characteristics of study population

The study was approved by the Institutional Ethical Boards of the National Institute of Biomedical Genomics, Kalyani, India and the SSKM Hospital, Kolkata. The study design is summarized in Fig. 1. A total of 551 participants were included in the study from two locations in West Bengal: Birbhum and SSKM hospital, Kolkata, with proper informed consent. Detailed medical history was noted by the hepatologist.

**Fig. 1:**
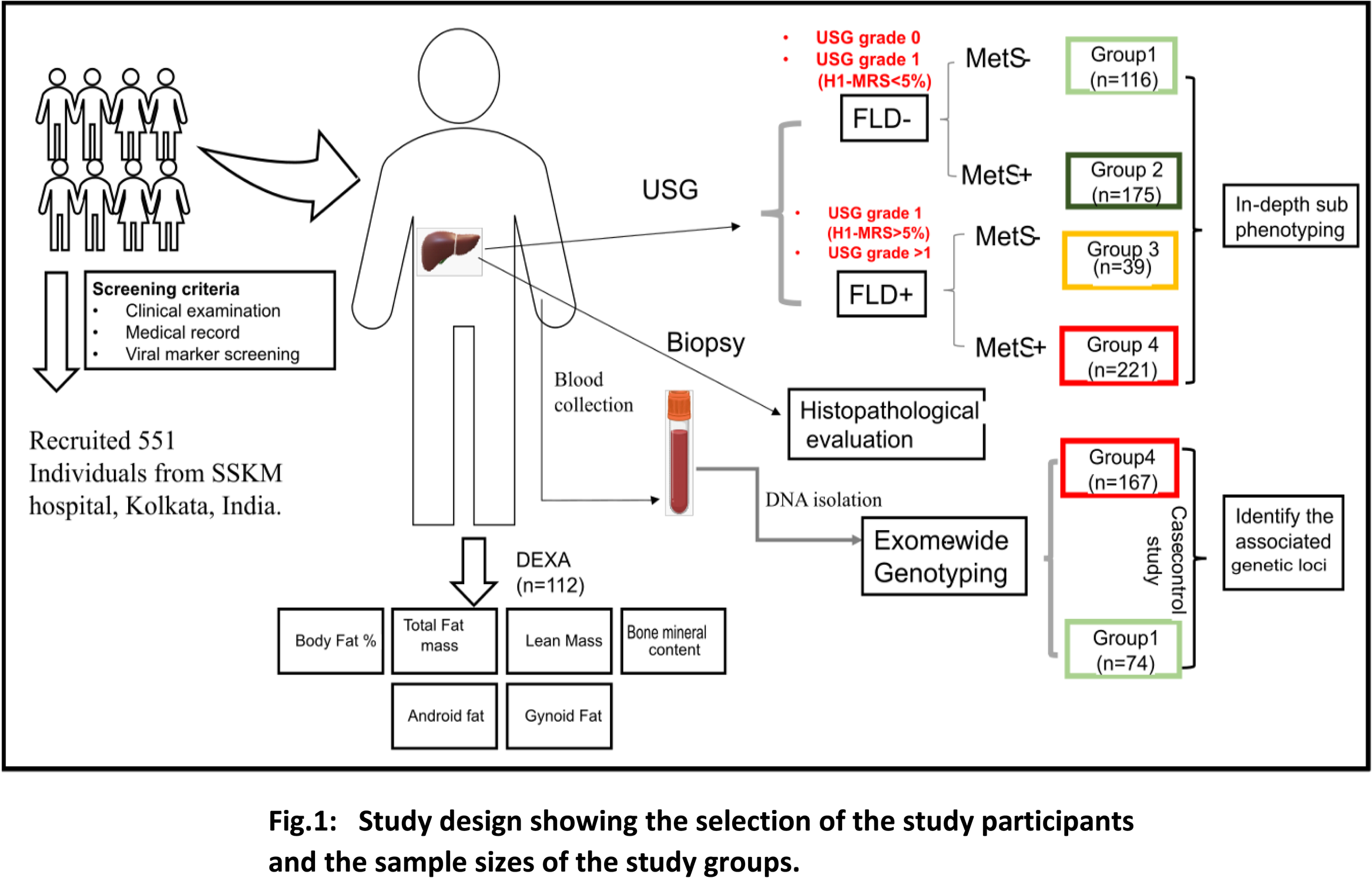
Study design showing the selection of the study participants and the sample sizes of the study groups.

After the initial screening based on the presence of viral markers, presence of fatty liver was examined through Ultrasonography (USG), performed by a single examiner [19]. Anthropometric measurements and blood biochemical investigations were performed on each study participants to assess the obesity status, lipid profile, liver damage, and metabolic profile. The study participants were sub-grouped into obese (with BMI >25kg/m2) and non-obese (with BMI <=25kg/m2). Homeostatic model assessment of insulin resistance (HOMA-IR) was calculated as previously described (Matthews et al., 1985), and a value of >1.64 was considered as significant IR. Liver stiffness measurement (LSM) data was done for all individuals using Fibroscan device (Echosens, France). Quantification of body composition was done using dual energy X-Ray absorptiometry (DEXA; GE Lunar Dexa BMD Machine). Detailed methology was mentioned in Supplementary methods.

### Characteristics of subgroups of patients

Due to limited repeatability of the USG based diagnosis, especially in case of low hepatic fat content, we further confirmed and quantified hepatic fat content using H_1_-MRS in a subgroup of patients (n=276) and observed the concordance of the results with that obtained from USG. The results showed that the diagnosis accuracy for Grade 0, Grade II and Grade III fatty liver were acceptable (83.63% for Grade 0, 90.2% for Grade II fatty liver and 100% for Grade III fatty liver). But for Grade I fatty liver the diagnosis accuracy was very low (53.6%) (Supplementary Fig. 1A). Thus, individuals with Grade I, II and III fatty liver under USG examination and/or >5% fat in liver under H_1_-MRS examination were suffering from fatty liver (FLD+). Individuals were considered as FLD-(no fat in liver), if absence of fatty liver (Grade 0) and/or <5% fat in liver under H_1_-MRS examination was detected. With these criteria a total of 260 FLD+ subjects and 291 FLD-subjects were finally included in the study (Supplementary Table 1, Supplementary Fig. 1: A, B)

Patients with both fatty liver and metabolic syndrome, were diagnosed based on the following criteria [7]:

1. Presence of fatty liver (USG grade 2 and 3)
2. Presence of adiposity (only for obese)
3. Presence of Type II Diabetes Mellitus
4. The presence of metabolic syndrome

The definition for identifying patients with each of the 4 above mentioned criteria are listed in Suppl. Fig. 1C. Based on the presence/absence of metabolic syndrome, the study cohort was divided into four subgroups – Group4_FLD+/MetS+_ (n=221), Group3_FLD+/MetS-_ (n=39), Group2_FLD-/MetS+_ (n=175) and Group1_FLD-/MetS-_ (n=116).

### Histopathological evaluation of disease severity

With informed consent liver tissue samples were collected from 179 patients through percutaneous needle biopsy, performed by a single clinician at SSKM Hospital. For histopathological evaluation, part of the tissue was preserved in 10% Formalin solution and embedded in paraffin cubes. Paraffinized samples were sectioned by microtome and stained with haematoxylin and eosin for morphological evaluation. Histological evaluation was performed according to the protocol mentioned in Brunt et al. 2011 by a single examiner. Disease severity was determined through NAFLD Activity Score (NAS) [20], [21], which is the sum of steatosis score, hepatocyte ballooning and lobular inflammation score. Degree of fibrosis was also determined by the fibrosis score of NASH-CRN criteria [21].

### Data curation and analysis

To study the differences in allele distribution at the selected loci between the Group4_FLD+/MetS +_ and Group1_FLD-/MetS-_, we used the genotype data from Chatterjee et al. 2021[18]. The data quality control and imputation were done as described in Chatterjee et al. 2021[18]. After quality control the final data included 34320 loci and 406 individuals. Among these 406 individuals, case-control association study was performed with 167 Group4_FLD+/ MetS+_ and 74 Group1_FLD-/ MetS-_ individuals. We have curated a list of SNPs from the NHGRI GWAS catalogue (https://www.genome.gov/) for the following traits –dyslipidemia, TG, HDL, LDL, and CRP. We hypothesized that the clinical phenotype of Group4_FLD+/ MetS+_ could have different genomic underpinnings of disease, compared to other subgroups. A list of 2123 SNPs were curated which were known to be associated with NAFLD or associated traits from NHGRI (www.genome.gov). Post quality control, further genetic association was performed on 2170 SNPs. We performed case-control study design to identify the variants associated with Group4_FLD+/ MetS+_ (n=167) compared to the Group 1_FLD-/ MetS-_ (n=74).

### Annotation of associated SNP

The associated SNP was mapped to nearby genes, using GRCh37/hg19 reference genome. The genomic location of the SNP and the annotated gene was visualized using UCSC Genome Browser (genome.ucsc.edu) and IGV. The details of the SNP including the effects of the nucleotide change were studied from dbSNP (www.ncbi.nlm.nih.gov/projects/SNP/snp_summary.cgi). The effects of the SNP on gene expression (single tissue eQTL) and the tissue specific gene expression among healthy tissues were observed using the GTEX consortium database (www.gtexportal.org/home/).

### Statistical analysis

Anthropometric variables and biochemical parameters between groups were compared using Wilcoxon-Singed Rank test as the distribution of most of the variables did not conform to ‘Normality’ assumptions. Comparisons for categorical variables were done using χ2 or Fishers exact test. Stepwise multiple logistic regression analysis was performed to identify significant predictors. We performed case-control association study to identify the association of SNPs with Group4_FLD+/MetS+_ compared to the Group1_FLD-/MetS-_. The analyses were done using PLINK v1.9. Benjamin-Hochberg FDR procedure was used for multiple testing corrections. All tests of significance were two-sided at a significance level of α= 0.05.

## Results

### Characteristics of the study population

In this study we recruited 551 individuals from two locations in the state of West Bengal: Birbhum and Kolkata (SSKM hospital). Among these 551 individuals, 288 (52.27%) were males and the mean age of the cohort was 41.75 ± 11.5 years (Supplementary Table 1). Our cohort consisted of both non-obese (n=277) individuals with a mean BMI of 21.42 ± 2.52 kg/m2 and obese individuals (n=274) with a mean BMI of 28.47 ± 3.11 kg/m2. .Out of 551 individuals, 31.76% had USG Grade I (n=175); 34.85% had Grade II (n=192), 4.7% Grade III (n=26) and 28.68% (n=158) were Grade 0 (no fat in liver) [22]. Type II diabetes (defined as FBG >=126 mg/dL and/or HbA1C > 6.4) was observed among 13.61% (n=75) of the study population, and 71.87% (n=396) had metabolic syndrome.

Clinico-pathological features of 260 FLD+ and 291 FLD-subjects were compared to identify the clinical risk factors for fatty liver in the study population (Supplementary Table 1). The groups did not differ in age (p-value = 0.58) and male to female ratio (p-value=0.08). FLD+ subjects had increased liver damage markers in the blood (ALT, AST and GGT) and increased liver stiffness measurement (LSM), thus indicating severity of liver disease. Measures of adiposity (weight, BMI, waist and hip circumference and skin fold thickness) were significantly higher among the FLD+ subjects. The FLD+ subjects had more severe diabetic profile (FBG, fasting insulin, HOMA-IR and HbA1c levels), than the FLD-subjects. When comparing the serum lipid profiles, the FLD+ subjects had significantly altered lipid profile with increased triglyceride, cholesterol, LDL and VLDL levels and decreased HDL levels than the FLD-subjects. The FLD+ subjects also had significantly increased serum CRP levels, thus indicating the presence of systemic inflammation [23].

### Fatty liver coupled with metabolic syndrome showed severe disease profile compared to other subgroups

Based on the presence/absence of metabolic syndrome (MeS), we subdivided our study cohort into four groups – Group4_FLD+/MetS+;_ (n=221), Group3_FLD+/MetS-;_ (n=39), Group2_FLD-/MetS+;_ (n=175) and Group1_FLD-/MetS-;_ (n=116) (Fig. 1). Comparing the clinicopathological features (Table 1), we observed that: (A) the measures of abdominal obesity (waist circumference) was highest in the Group4_FLD-/MetS+_ (mean= 93.68cm), followed by in Group3_FLD+/MetS-_ (mean= 90.54cm) (p-value <0.01); (B) As expected, Group4_FLD+/MetS+_ patients had more severe diabetic profile and higher HOMA-IR than Group3_FLD+/MetS-_. However, Group4_FLD+/MetS+_ patients also had significantly severe diabetic profile than Group2_FLD-/MetS+_ (only MeS) individuals; (C) Both the Group4_FLD+/MetS+_ and Group3_FLD+/MetS-_ showed higher liver damage markers with significantly higher ALT and AST levels, than the Group2_FLD-/MetS+_ and Group1_FLD-/MetS-_. (D)We observed CRP levels were significantly higher in Group4_FLD+/MetS+_ individuals compared to other groups. (E) Among the patients with LSM values beyond the critical threshold (>7KPa), majority were Group4_FLD+/MetS+_ (61.63% in Group4, 0.64% in Group3 and 4.31% in Group 2) (Table 1).

**Table 1:**
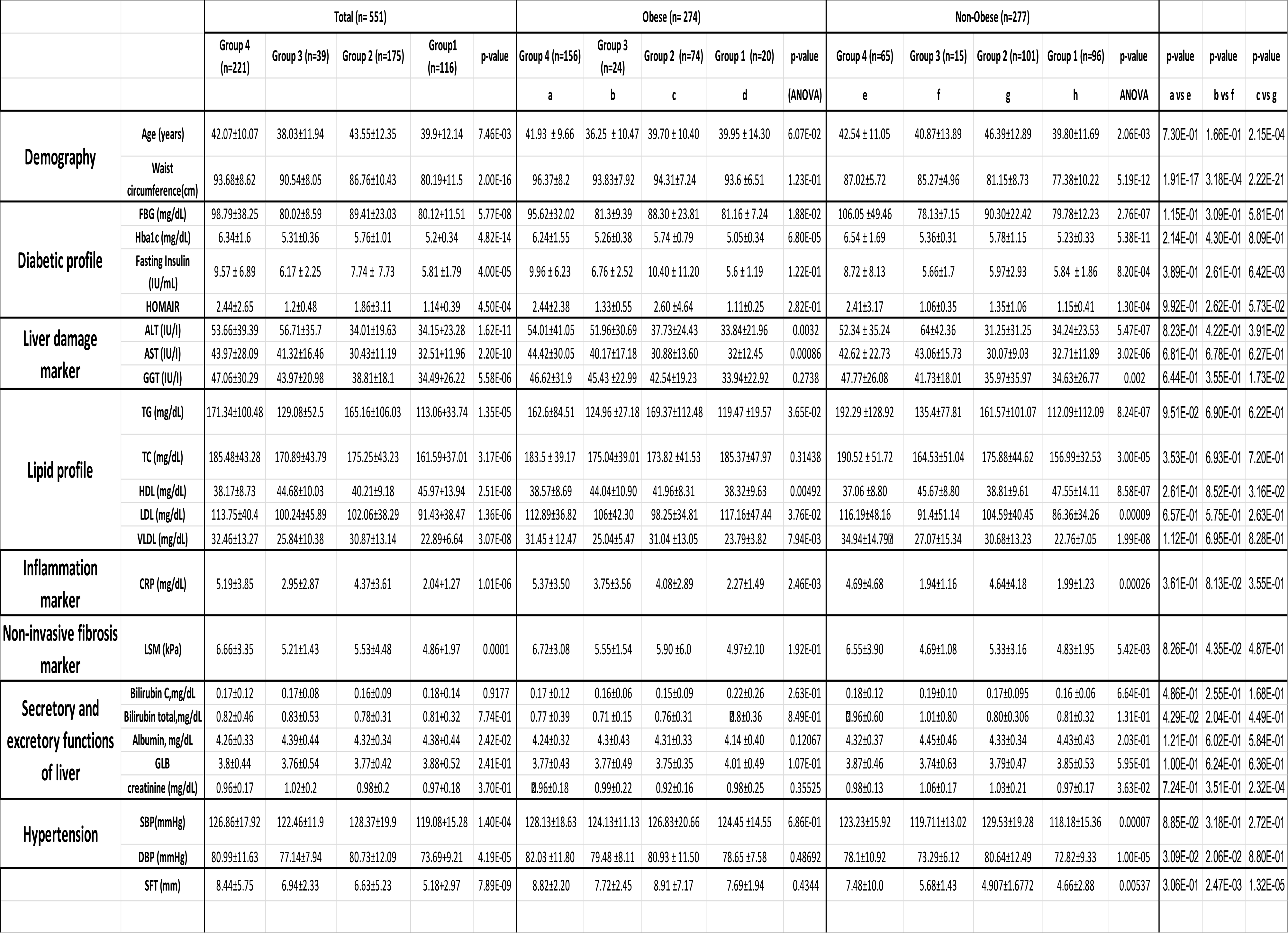
Pathophysiological risk factors associated with metabolic syndrome across study groups.

Group4_FLD+/MetS+_ individuals had significantly higher CRP (mean=5.19 mg/dL in Group4_FLD+/MetS+_ and 2.04 mg/dL in Group1_FLD-/MetS-_), LSM (mean=4.86 kPa in Group1_FLD-/MetS-_ and 6.67 kPa in Group4_FLD+/MetS+_, TG mean=171.34 mg/dL in Group4_FLD+/MetS+_ and 113.06 mg/dL in Group1_FLD-/MetS-_) and lower HDL level (mean=45.97 mg/dL in Group1_FLD-/MetS-_ and 38.17 in Group4_FLD+/MetS+_) after adjusting for the effects of other covariates (Fig. 2:A-D). The Group4_FLD+/MetS+_ individuals had also significantly higher LSM (mean= 5.21 kPa in Group3_FLD+/MetS-_ and 6.66 kPa in Group4_FLD+/MetS+_) and lower HDL compared to Group3_FLD+/MetS-_(mean= 44.68 mg/dL in Group3_FLD+/MetS-_ and 38.17 in Group4_FLD+/MetS+_) (Fig. 2: E-F). Interestingly, Group3_FLD+/MetS-_ had higher serum creatinine (mean=1.02 mg/dL in Group3_FLD+/MetS-_ and mean=0.96 mg/dL in Group4_FLD+/MetS+_ than Group4_FLD+/MetS+_, after adjusting the effects of other covariates (age, obesity, lipid profiles and liver damage markers). Finally, our results also showed that the Group4_FLD+/MetS+_ individuals had higher BMI (mean=27.19 Kg/m2 in Group4_FLD+/MetS+_ and 24.15 Kg/m2 in Group2_FLD-/MetS+_) and ALT (mean=53.66 in Group4_FLD+/MetS+_ and 34.01 in Group2_FLD-/MetS+_ than Group2_FLD-/MetS+_) (Fig. 2:G-H). These observations suggested that Group4_FLD+/MetS+_ individuals might have higher obesity and more severe disease profile than Group3_FLD+/MetS-_ and Group2_FLD-/MetS+_.

**Fig. 2:**
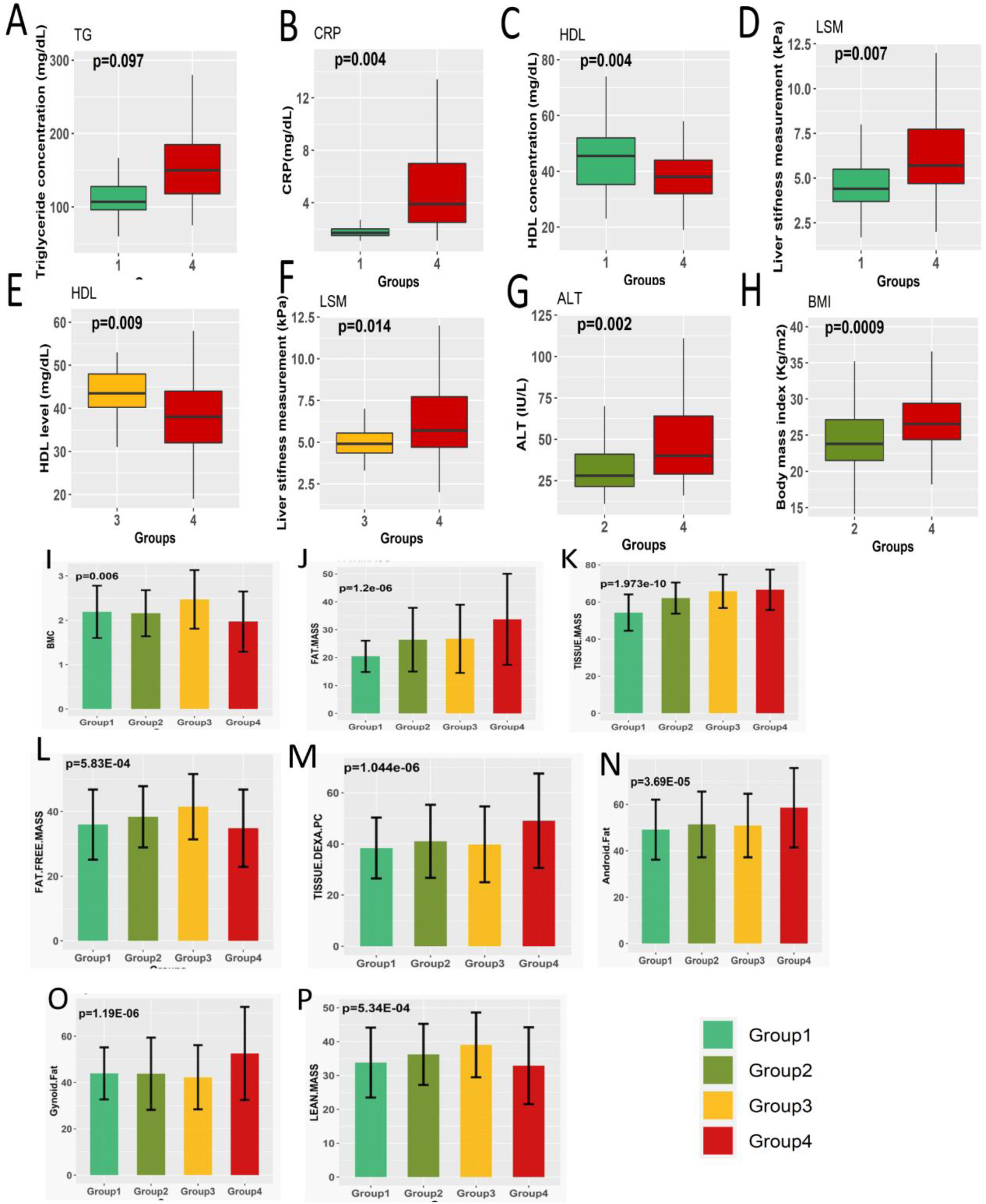
Differences in the body composition using anthropometry and DEXA and blood biochemical parameters among the four subgroups: Group4_FLD+/MetS+_ vs. Group1_FLD-/MetS-_ (A-D), Group4_FLD+/MetS+_ vs. Group3_FLD+/MetS-_ (E-F), and Group4_FLD+/MetS+_ vs. Group2_FLD-/MetS+_ (G, H). p-values depicted level of significance, adjusting for the effects of other covariates. **DEXA parameters** – (I) Bone mineral content, (J) fat mass, (K)tissue mass, (L) fat free mass, (M) total body fat (%), (N) Android fat %, (O) Gynoid fat %, and (P) lean mass.

### Bone mineral content was observed lowest among fatty liver patients coupled with metabolic syndrome

Analysis of body fat, muscle composition and bone mineral content were done using DEXA (Supplementary Table 2). Out of 551 study participants, DEXA was performed for 112 study participants (Fig. 1; Group4_FLD+/MetS+_, n=60; Group3_FLD+/MetS-_, n=14; Group2_FLD-/MetS+_, n=31 and Group1_FLD-/MetS-_, n=7). Group4 _FLD+/MetS+_ had significantly higher percentages of total body fat (TBF) (Fig. 2.M), android and gynoid fat, and tissue mass compared to the rest three groups, after adjusting for age and gender (Supplementary Table 2; Fig. 2:K-O). Interestingly, Group4_FLD+/MetS+_ had the lowest bone mineral content (BMC) compared to the other three groups (Fig. 2.I). Group3_FLD+/MetS-_ individuals had lower TBF, higher tissue mass and higher BMC compared to Group2_FLD-/MetS+_ (Suppl. table 2) (Fig. 2: I, K, M).

### Presence of metabolic syndrome is associated with NASH and liver damage among the fatty liver patients

For a subgroup of individuals (n=168), histopathological evaluation was performed to assess the disease severity. Among them, 112 were Group4_FLD+/MetS+_, 21 were Group3_FLD+/MetS-_ and 35 were Group2_FLD-/MetS+_ (Fig. 3). Our results showed that Group4_FLD+/MetS+_ had more severe disease phenotype compared to Group3_FLD+/MetS-_ and Group2_FLD-/MetS+_; that is, 41.07% of Group4_FLD+/MetS+_ had NASH (NAS >3), compared to 19.05% in Group3_FLD+/MetS-_ and 5.71% in Group2_FAT/MetS+_ (Fig. 3.F). Proportions of each of the features of the NAS, including the steatosis score (Fig. 3.A), hepatocyte ballooning score (Fig. 3.B) and inflammation (Fig. 3:C-D), were highest in the Group4_FLD+/MetS+_ compared to any other groups. Severe fibrosis (fibrosis score >=3) was also observed more in Group4_FLD+/MetS+_ (6.25%) compared to Group3_FLD+/MetS-_ (4.76%) and Group2_FLD-/MetS+_ (2.85%) (Fig. 3.E). This evidence is further supported by highest mean LSM values in Group4_FLD+/MetS+_, compared to the other groups (Table 1).

**Fig. 3:**
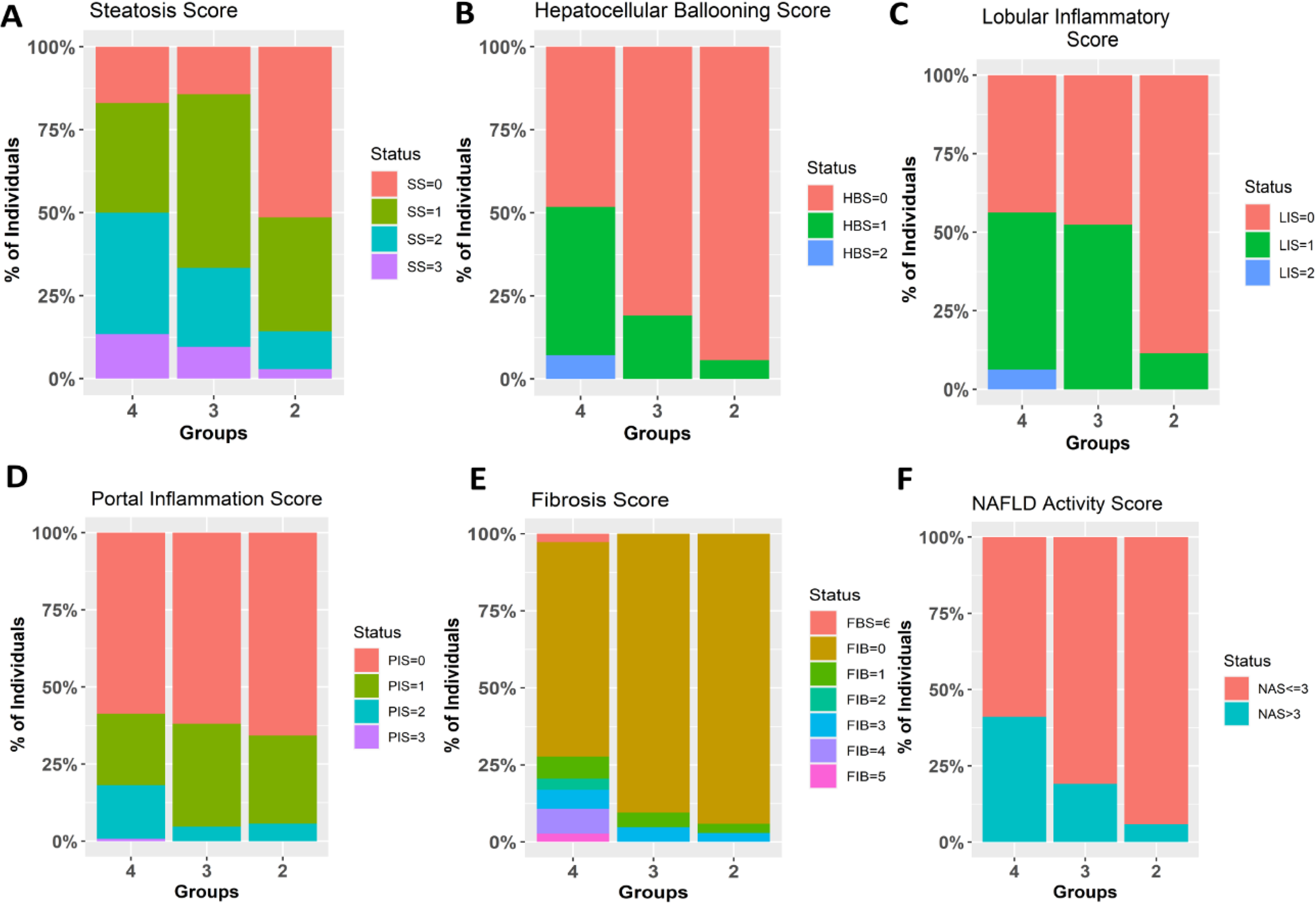
Distribution of patients with different degrees of disease severity across the four study groups: A) Steatosis score B) Hepatocellular ballooning score C) Lobular inflammatory score D) Portal inflammation score E) Fibrosis score F) NAFLD activity score

### Metabolic syndrome is associated with disease severity, irrespective of the obesity status

Our previous study along with other studies in the last decade have shown that in Asian and South-east Asian populations non-alcoholic fatty liver is associated with the “lean” phenotype [24]. Obesity is highly associated with MeS; however deranged metabolic profiles are also observed among individuals without any sign of obesity [25]. In our cohort, we found 49.73% (n=274) individuals were obese and 50.27% (n=277) individuals were non-obese. Abdominal obesity was present in 80.04% (n=441) of the study cohort and 23.4% (n=172; males=130, females=42) of the non-obese cohort. As expected, among the obese and non-obese population, the Group4_FLD+/MetS+_ individuals had significantly higher FBG, HbA1c, AST, ALT, VLDL, CRP and LSM than Group3_FLD+/MetS-_ and Group2_FLD-/MetS+_. HDL was lowest in Group4_FLD+/MetS+_ than others three groups, both among the obese and the non-obese (Table 1).

Non-obese Group4_FLD+/MetS+_ had significantly higher waist circumferences (87.02±5.72), than the rest of the non-obese subgroups (Table 1). Fasting insulin level was significantly higher in Group4_FLD+/MetS+_ (8.72 ± 8.13) than Group3_FLD+/MetS-_ (5.66±1.7) and Group2_FLD-/MetS+_ (5.97±2.93) and HOMA-IR was significantly increased in Group4_FLD+/MetS+_ (2.41±3.17) than other groups (Group3_FLD+/MetS-_ (1.06±0.35), Group2_FLD-/MetS+_ (1.35±1.06)). Among non-obese Group2_FLD-/MetS+_ individuals who had only metabolic syndrome, we found the diabetic profile was comparatively significantly higher than non-obese Group3_FLD+/MetS-_ (Table 1). The non-obese Group4_FLD+/MetS+_ also had more severe liver damage markers (ALT, AST, GGT, LSM) and dysregulated lipid profiles than other three groups.

To study the effects of obesity and metabolic syndrome exclusively on fatty liver development, we had compared the clinico-pathological features of obese Group3_FLD+/MetS-_ (n=24) with non-obese Group2_FLD-/MetS+_ individuals (n=101) (Table 2). Diabetic profile including the FBG, HOMA-IR and HbA1c levels were higher in non-obese Group2_FLD-/MetS+_. Non-obese Group2_FLD-/MetS+_ also had higher serum TG and lower HDL levels. The levels of CRP were also higher among the non-obese Group2_FLD-/MetS+_ patients than the obese Group3_FLD+/MetS-_ patients (Table 2).

**Table 2:** Comparison of pathophysiological risk factors between obese Group3_FLD+/MetS-_ individuals with non-obese Group2_FLD-/MetS+_.

|  |  |  |  |
| --- | --- | --- | --- |
|  | Obese | Non-Ob | p-value |
|  | Group 3 (n=24) | Group 2 (n=101) |  |
| Age (years) | 36.25 ± 10.47 | 46.39±12.89 | **2.58E-05** |
| Waist circumference(cm) | 93.83±7.92 | 81.15±8.73 | **1.21E-07** |
| FBG (mg/dL) | 81.3±9.39 | 90.30±22.42 | **0.004** |
| Hba1c (mg/dL) | 5.27±0.39 | 5.78±1.15 | **0.001** |
| Fasting Insulin (IU/mL) | 6.76 ± 2.52 | 5.97±2.93 | 0.395 |
| HOMAIR | 1.33±0.55 | 1.35±1.06 | 0.767 |
| ALT (IU/I) | 51.96±30.69 | 31.25±31.25 | **0.004** |
| AST (IU/I) | 40.18±17.18 | 30.07±9.03 | **0.009** |
| GGT (IU/I) | 45.44±22.99 | 35.97±35.9673 | 0.067 |
| TG (mg/dL) | 124.96 ±27.18 | 161.57±101.07 | **0.002** |
| TC (mg/dL) | 175.04±39.01 | 175.88±44.62 | 0.714 |
| HDL (mg/dL) | 44.04±10.90 | 38.81±9.61 | **0.031** |
| LDL (mg/dL) | 106±42.30 | 104.59±40.45 | 0.870 |
| VLDL (mg/dL) | 25.04±5.47 | 30.68±13.23 | **0.002** |
| CRP (mg/dL) | 3.75 ±3.56 | 4.64±4.18 | 0.404 |
| LSM (kPa) | 5.55±1.54 | 5.33±3.15 | 0.538 |
| Bilirubin C,mg/dL | 0.16±0.057 | 0.17±0.095 | 0.317 |
| Bilirubin total,mg/dL | 0.71±0.15 | 0.80±0.31 | 0.067 |
| Albumin, mg/dL | 4.34±0.43 | 4.33±0.34 | 0.879 |
| GLB | 3.78±0.49 | 3.79±0.47 | 0.961 |
| creatinine (mg/dL) | 0.99±0.22 | 1.03±0.21 | 0.510 |
| SBP(mmHg) | 124.13±11.13 | 129.53±19.28 | 0.095 |
| DBP (mmHg) | 79.48 ±8.11 | 80.64±12.49 | 0.632 |
| SFT (mm) | 7.73±2.45 | 4.90±1.68 | **8.50E-06** |

### Genetic predisposition of fatty liver coupled with metabolic syndrome

To study the genetic predisposition to fatty liver coupled with metabolic syndrome, we took a candidate gene approach and identified a list of SNPs which were previously reported to be associated with dyslipidaemia, HDL, TG, LDL, CRP (EFO_0004530, EFO_0009946, EFO_0003095, MONDO_0004790, EFO_0007805, EFO_0000195, EFO_0006890). Among 392 curated SNPs, significant associations with Group4_FLD+/MetS+_ were observed for 21 SNPs (p-value <0.05) (Table 4). No SNPs showed significant association after multiple testing corrections, possibly due to small size. Among the 21 SNPs, four SNPs were missense nucleotide changes with odds ratios of the risk alleles >1; namely, rs3761472-G (*SAMM50,* OR=2.9 (2.0-4.1)), rs738409-G (*PNPLA3,* OR=2.8 (1.9-4.07)), rs58542926-A (*TM6SF2,* OR=2.7(1.9-3.9)), and rs35665085-A (*CECR5,* OR=2.7 (1.9-3.9)) (Supplementary Table 3).

Among the rest, three SNPs had significantly higher odds ratios – rs471364-G (*TTC39B,* OR=3.1 (2.1-4.5)), rs2800-G (*SLC9A9,* OR=3.1 (2-4.5)), rs7200543-A *(PDXDC1,* OR=2.1 (1.1-4.8)). Comparing with the allele frequencies of the five populations in the 1000G database, we observed that the risk allele frequencies of associated SNPs in our healthy cohort were similar to those in the Asian population (Supplementary Table 3). The nearby (<0.5Mb) SNPs were found to be in high linkage disequilibrium (>0.6) with the sentinel SNPs in the South-East Asian population cohort of the 1000G data (Fig. 4,5). Additionally, we also observed that for SNPs – rs1532085 (*CECR5*), rs58542926 (*TM6SF2*), rs2800 (*SLC9A9*) – the risk allele frequencies were highest among the Group4_FLD+/MetS+_ patients than the other two groups (Group3_FLD+/MetS-_ and Group2_FLD-/MetS+_). In the past decade, multiple studies have clearly established the involvement of variants in *PNPLA3, SAMM50, TM6SF2* genes in fatty liver pathogenesis [18], [26]. Like previous finding, our results also showed higher risk allele frequencies for rs738409, rs3761472 and rs58542926 SNPs among Group4_FLD+/MetS+_ compared to the Group1_FLD-/MetS-_ (Supplementary Fig. 2-4).

**Fig. 4:**
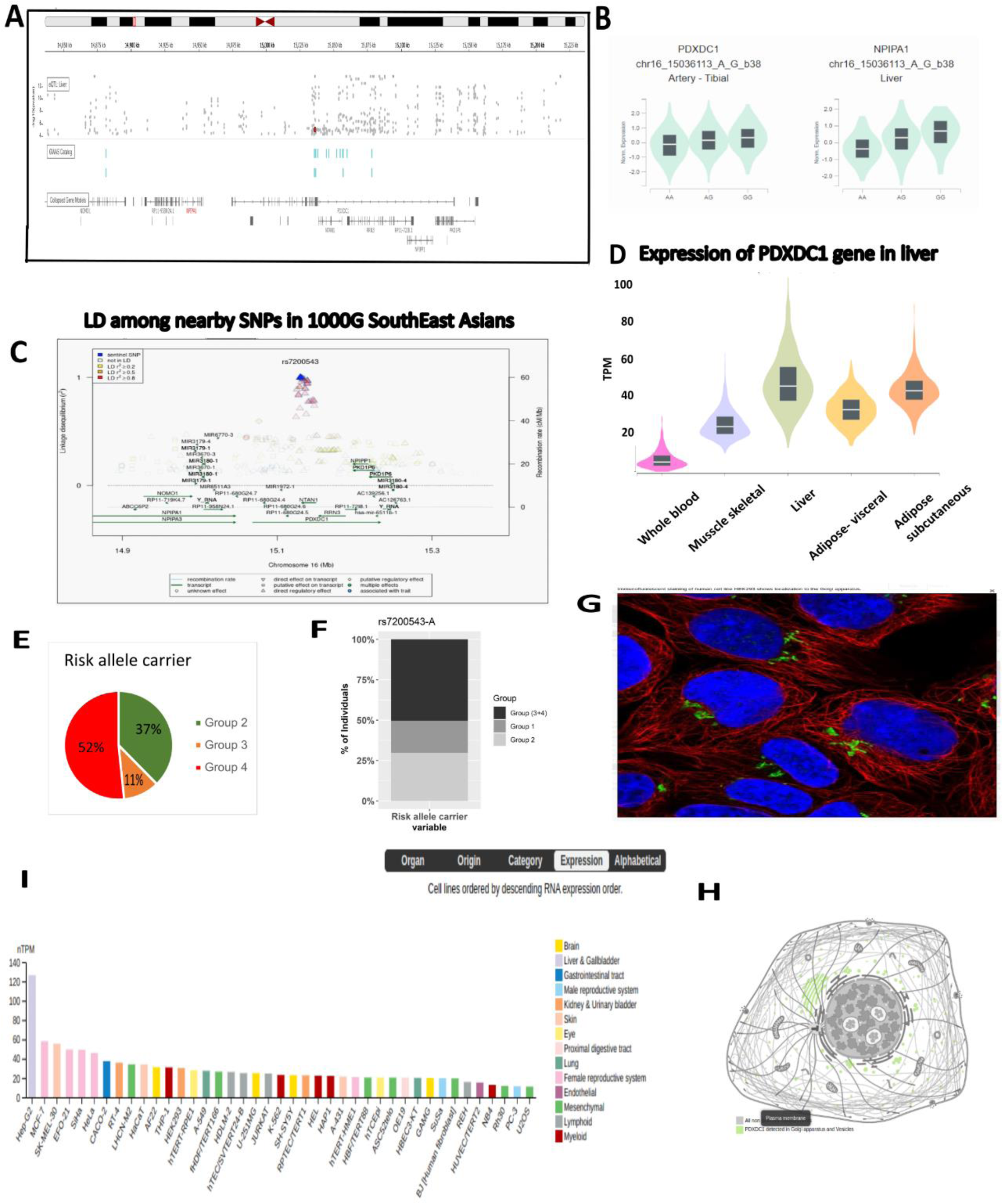
Association of rs72400543 in PDXDC1: (A) Genomic regions, transcripts, and products from dbSNP Short Genetic Variations (hg19). (B) Liver tissue specific effect of the SNP on *NPIPA1* and *PDXDC1* gene expressions. (C) Linkage Disequilibrium plot from SNiPA showing high LD among the nearby SNPs and rs72400543 in south-east Asian population. (D) Tissue specific expression of *PDXDC1* gene expression from GTEx portal for five different tissue types – adipose, skeletal and liver. (E-F) Percentage of risk allele carrier in Group4_FLD+/MetS+,_ Group3_FLD+/MetS-_ and Group2_FLD-/MetS+._ (G) Subcellular location of *PDXDC1* protein in golgi-apparatus and vesicles based on immunofluorescent analysis from Human Protein Atlas. (H) Schematic of intracellular location of PDXDC1 gene and protein from Human Protein Atlas. (I) Expression plot of PDXDC1 gene across the different cell lines from Human Protein Atlas.

**Fig. 5:**
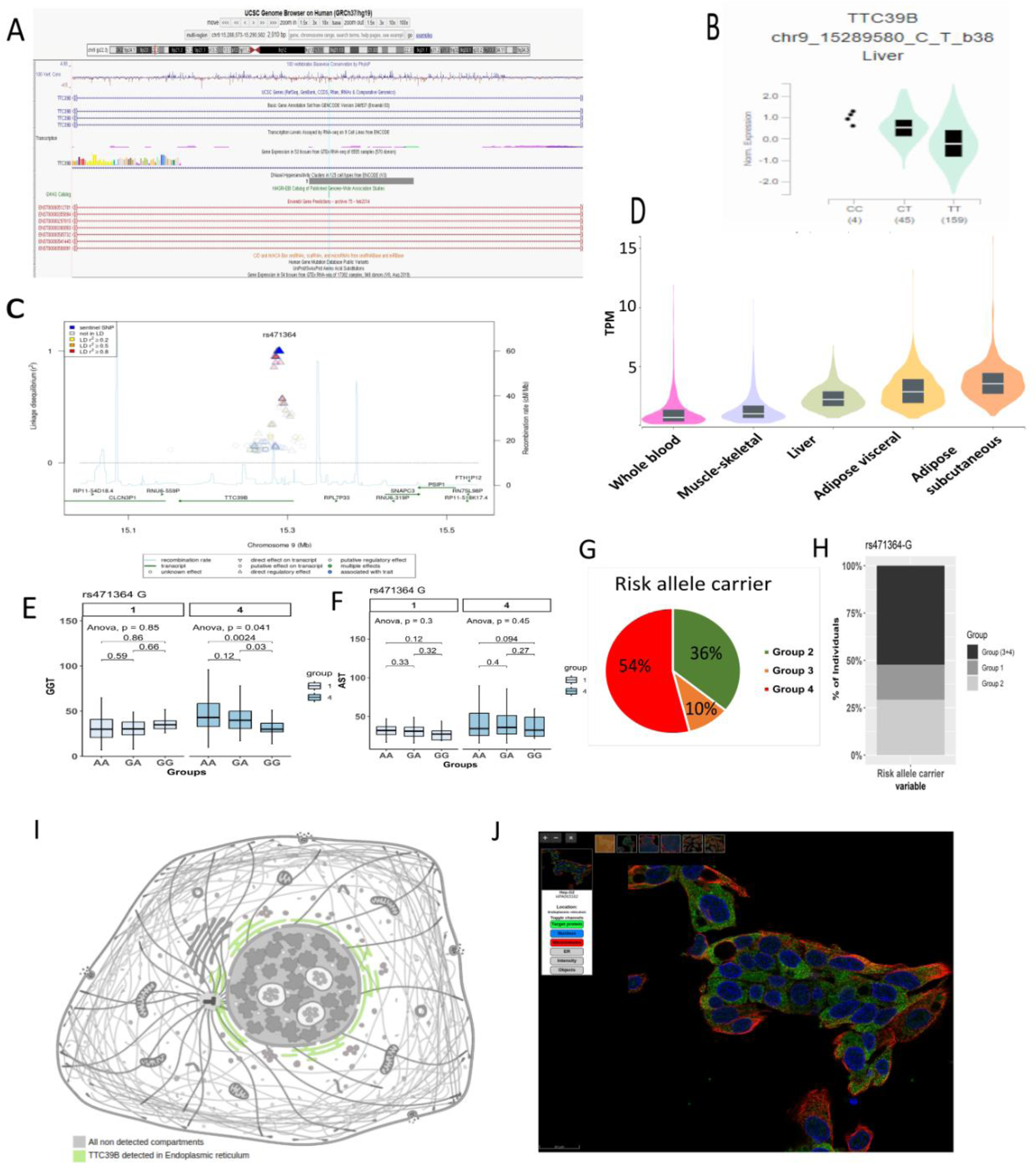
Association of rs471364 in *TTC39B*. (A) Genomic regions, transcripts, and products from dbSNP Short Genetic Variations (hg19). (B) Liver tissue specific effect of the SNP on gene *TTC39B* gene expression. (C) Linkage Disequilibrium plot from SNiPA showing high LD among the nearby SNPs and rs471364 in south-east Asian population. (D) Tissue specific expression of *TTC39B* gene expression from GTEx portal for five different tissue types – adipose, skeletal and liver. (E-F) Distribution of liver damage markers among different genotypes. (G-H) Percentage of risk allele carrier in Group4_FLD+/MetS+,_ Group3_FLD+/MetS-,_ Group2_FLD-/MetS+_ and Group1_FLD-/MetS-_) Schematic diagram of intracellular location of TTC39B protein from THE HUMAN PROTEIN ATLAS. J) Subcellular location of TTC39B in endoplasmic reticulum based on immunofluorescent analysis from Human Protein Atlas.

Among the other associated SNPs, rs471364-G (*TTC39B*) and rs7200543 (*PDXDC1*) were investigated further. We searched the GTEX database for gene expression and found that *PDXDC1* and *TTC39B* genes were highly expressed in liver and adipose tissues compared to blood (Fig. 4,5). For the homozygous risk allele carriers, we observed significantly higher liver damage markers including GGT, ALT and AST and low albumin (Fig. 4,5). We also found that the risk allele carriers of the two SNPs had significantly low bone mineral content than the others (Fig. 4,5). To understand the effects on the variants on gene expression, we searched GTEX eQTL database (https://gtexportal.org/home/tissue/). Both the SNPs were reported to have liver specific eQTL effect on gene expression – rs7200543 for *NPIPA1* and rs471364 for *TTC39B*. The SNP rs471364-T allele was associated with reduced expression of *TTC39B* gene and rs7200543-A was associated with reduced expression of *NPIPA1* gene in the liver tissues (Fig. 4,5). The risk alleles at these two loci were also associated with higher albumin, lower bone mineral content and higher liver damage markers (Fig. 4,5).

## Discussion

NAFLD is a chronic liver disease with its pathologic state NASH, characterized by inflammation and fibrosis, is the potentially progressive form of NAFLD [20]. Studies in the past decade have identified that among the risk factors associated with NASH development, metabolic syndrome (MetS) is an important one [20]. However, a major criticism was that fatty liver disease can also affect individuals in absence of MetS, especially in countries where fatty liver disease is observed among the non-obese populations [27]–[29]. Here, we report the first study from India, in which we have characterized the pathophysiological features and genetic predisposition of NAFLD patients with MetS, compared to patients with only fatty liver and with only MetS.

Consistent with previous studies, stating that majority (81.92%) of the non-alcoholic fatty liver patients had metabolic syndrome, we observed a prevalence of 71.87%[8], [30], [31]. 7.08% of the patients had only fatty liver (Group3_FLD+/MetS-_) and 31.76% suffered from only MetS (Group2_FLD-/MetS+_) without any fatty liver [8]. As expected, the percentages of (Group4_FLD+/MetS+_) patients were higher among the obese (BMI >25 kg/m^2^; 56.93%) compared to the non-obese (23.47%), thus reiterating the fact that obese individuals are prone to severe MetS [13], [32]. This observation is like a previous report by Chen et al., who showed the prevalence of MetS among lean NAFLD patients is 25.3% [16]. De-regulated diabetic profile and lipid profile were observed more in the Group4_FLD+/MetS+_ compared to Group3_FLD+/MetS-_ or Group2_FLD-/MetS+_ [8], [16]. A previous study had reported that metabolically unhealthy individuals carry higher risk of fatty liver disease than healthy individuals [16]. It is known that MetS can aggravate prognosis of liver diseases [33] and contributes to higher risk of end-stage liver disease (cirrhosis and liver cancer) [34]. We confirmed that in our study cohort, Group4_FLD+/MetS+_ patients had severe liver diseases with increased steatosis score, hepatocyte ballooning, portal and lobular inflammation and NAFLD activity score along with liver damage markers. In fact, the presence of MetS could be a more significant risk factor than the presence of only mild steatosis, as observed by the higher percentage of individuals with severe disease among the Group2_FLD-/MetS+_ patients compared to Group3_FLD+/MetS-_ patients (Fig. 3). Previous studies have shown that 53.7% of fatty liver patients and 17.3% patients with metabolic syndrome had de-regulation of liver metabolic functions [33]. We observed that Group4_FLD+/MetS+_ patients had significantly lower serum albumin levels and higher bilirubin levels (both total and conjugated) than the other groups. Low albumin level indicates poor synthetic function of liver while higher bilirubin indicates poor excretory function of the liver [35]. Low albumin is associated with several liver diseases including NAFLD [35].

Bone mineral content (BMC) and lean body mass are indications of good skeletal health. Past studies have shown that poor skeletal health was associated with lower BMC [36]. Additionally, hyperglycaemia and adiposity may be related to reduction in bone mineral density [37], [38]. Low HDL levels and an inflammatory microenvironment affect the differentiation and function of osteoblasts [39]. In our study, we observed significant reduction of BMC and lean body mass, while increased fat mass among the Group4_FLD+/MetS+_ individuals compared to other groups, suggesting association of poor skeletal health among the fatty liver patients with metabolic syndrome.

We showed that Group4_FLD+/MetS+_ patients had significantly higher abdominal obesity and skin fold thickness than both Group3_FLD+/MetS-_ and Group2_FLD-/MetS+_. Past studies have shown that visceral adiposity is more prevalent among the Asian and South-Asian populations [40]. Increased visceral obesity was previously shown to predict liver fibrosis among the non-alcoholic individuals [41], [42]. Our data showed that non-obese Group4_FLD+/MetS+_ had lesser systemic metabolic derangements than the obese Group4_FLD+/MetS+_ patients [16]. Non-obese Group2_FLD-/MetS+_ individuals had lower HDL than the obese fatty liver patients (Group3_FLD+/MetS-_). Again, diabetic profile (FBG, HbA1C) and lipid profile such as triglyceride and VLDL were significantly higher in non-obese MeS individuals (Group2_FLD-/MetS+_), compared to obese fatty liver patients (Group3_FLD+/MetS-_). Additionally, levels of systemic inflammation marker (CRP), blood pressure (SBP, DBP), bilirubin (total bilirubin, conjugated bilirubin) and creatinine were significantly higher in non-obese Group2_FAT/MetS+_ compared with obese Group3_FLD+/MetS-_ individuals. These observations clearly indicated that the presence of metabolic syndrome could be a surrogate of liver damage.

Genetic association study identified six SNPs to be associated with NAFLD patients with metabolic syndrome in our study population – among them, three SNPs were previously reported to be robustly associated with fatty liver disease (rs738409-G, rs3761472-G and rs58542926-A) [18], [26]. Additionally, we also observed association with rs471364-G(*TTC39B*), rs7200543-A (*PDXDAC1*) and rs35665085-A (*CECR5/HDHD5*). Among the three novel associated SNPs, rs471364-G is a silent nucleotide change in *TTC39B* gene. When searched in the GTEX database, the rs471364-G was associated with increased expression of *TTC39B* gene in the liver. Previously, the *TTC39B* gene was found to be associated with HDL in European population [43]. *TTC39B* (Tetratricopeptide Repeat Domain 39B) regulates HDL metabolism by promoting the ubiquitination and degradation of the oxysterol’s receptors LXR [44]. Using Protein Atlas Database, the subcellular location of the *TTC39B* protein was found to be in the Endoplasmic reticulum and is likely involved in lipid metabolism and transport processes, like *PDXDC1*. The SNP rs35665085-A is a missense nucleotide change in *CECR5* gene (Missense_T149M). Previously, *CECR5* gene was found to be associated with higher triglyceride and body weight in European population [45]. Finally, rs7200543-A in *PDXDC1* gene is a synonymous nucleotide change (L735L). Interestingly, from the GTEX database rs7200543-A was found to be an eQTL of *NPIPA1* and *PDXDC1* genes and the risk allele is associated with decrease in *PDXDC1 and NPIPA1* expressions. A recent study had shown that higher expression of *PDXDC1* gene is related to higher Bone Mineral Density (BMD) and lower Polyunsaturated fatty acid (PUFA) levels, which may imply a protective effect of the gene under metabolic syndrome [46]. We further showed that the risk allele careers for the associated SNPs-rs7200543-A, rs3761472-G, rs738409-G had significantly lower BMC compared to non-risk allele careers [36], [47]–[50].

## Conclusions

In this study, we performed in-depth sub-phenotyping of the fatty liver disease in the presence/absence of metabolic syndrome (MeS) in a cohort of Indian patients. MeSwas prevalent among the NAFLD patients (71.87%), especially among the obese; the data was comparable to the only two previous studies from South-East Asian population. Irrespective of the obesity status, NAFLD patients with MeS had more severe disease profile, higher abdominal obesity and lower bone mineral content than either of patients with only fatty liver or with only MeS. Non-obese individuals with MeS had higher systemic metabolic derangements than obese fattyliver patients without MeS. Finally, we also showed the association of known SNPs in PNPLA3 and SAMM50 genes along with novel SNPs in PDXDC1 and TTC39B genes with fatty liver disease coupled with metabolic syndrome.

### Conflict of interests

The authors declare no potential conflict of interests.

### Financial support

Department of Biotechnology, Government of India. Intramural funding.

### Data availability

All the raw data and processed data are available upon reasonable request. **Acknowledgements:**

The authors would like to thank all the study participants who consented for the study.

## Data Availability

All data produced in the present study (including the raw data, processed data and the metadata) are available upon reasonable request to the authors.

## Notes

### Competing Interest Statement

The authors have declared no competing interest.

### Author Declarations

The study was approved by the Institutional Ethical Boards of the National Institute of Biomedical Genomics, Kalyani, India and the SSKM Hospital, Kolkata (Ethical approval number:NIBMG/2012/1/6)

