## Supplementary Files for "Severe liver damage, low bone mineral content, and PDXDC1/TTC39B gene variations linked to NAFLD with metabolic syndrome"

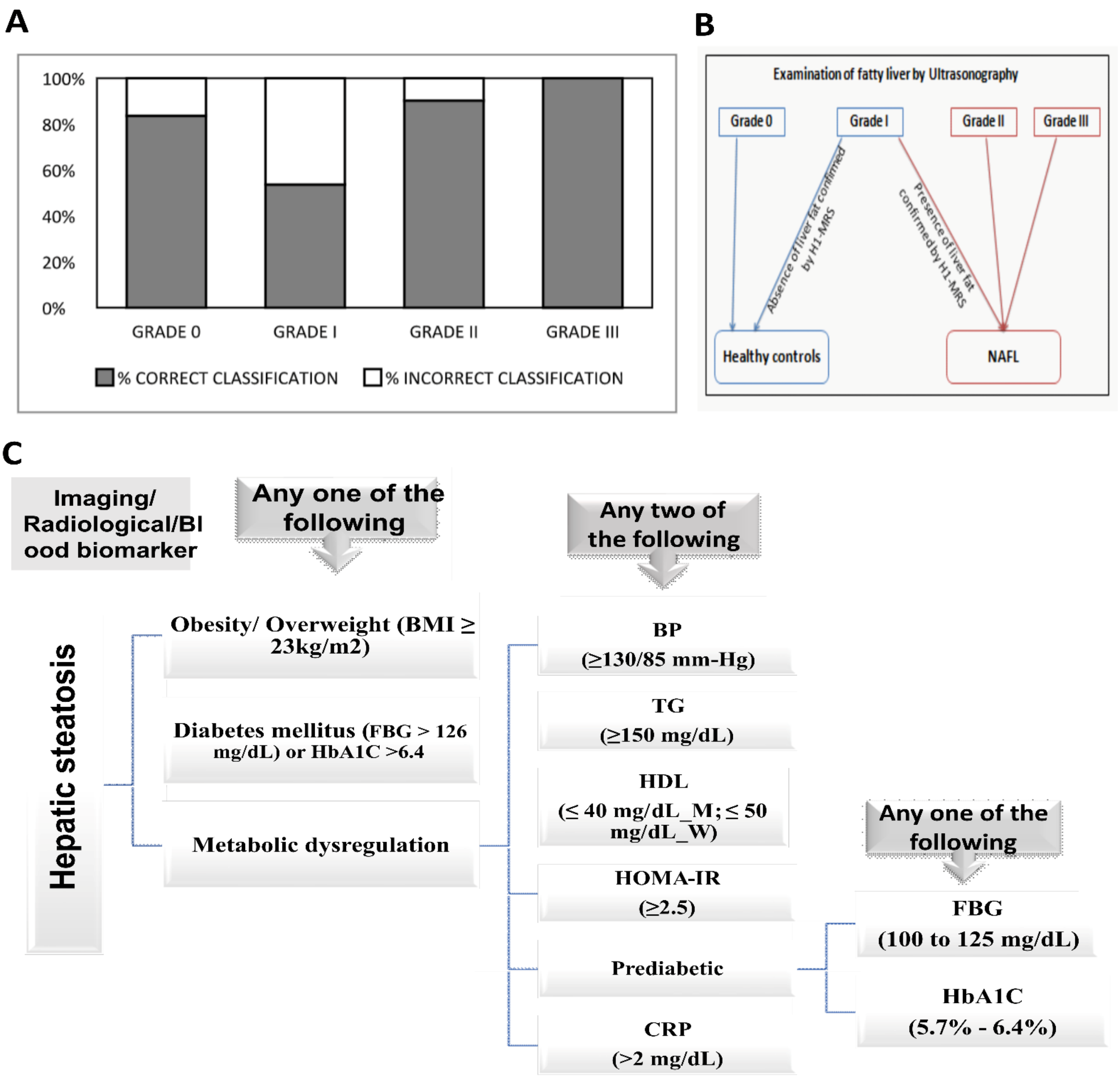

**Supplementary figure 1:**

A) Concordance of diagnosis of fatty liver performed by both MRS and ultrasonography; B) Diagnostic definitions of healthy and fatty liver patients, followed in our study; C) Diagnostic definition of Group4 FLD+/MeS+

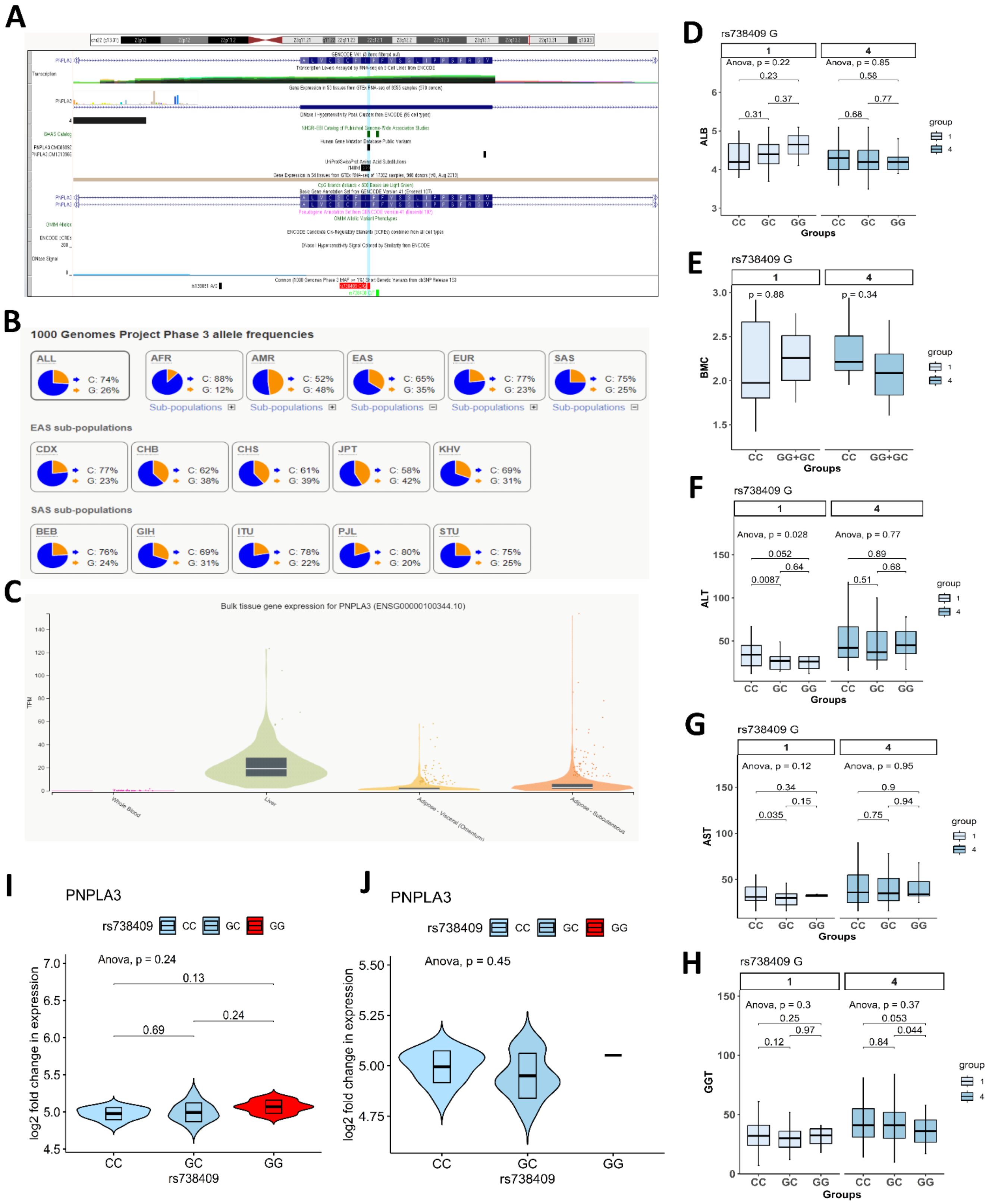

**Supplementary figure 2: Details of rs738409**

A) Genomic annotation of rs738409 in PNPLA3 gene from dbSNP Short Genetic Variations (hg19);B) 1000 Genomes Project Phase 3 allele frequencies from ensembl.org ;C) Gene expression from GTEx portal for five different tissue types – adipose, skeletal and liver; D-H) Distribution of liver damage markers and bone mineral content among different genotypes;I- J) violin plots showing the effects of eQTL on PNPLA3 gene expression in our study cohort.

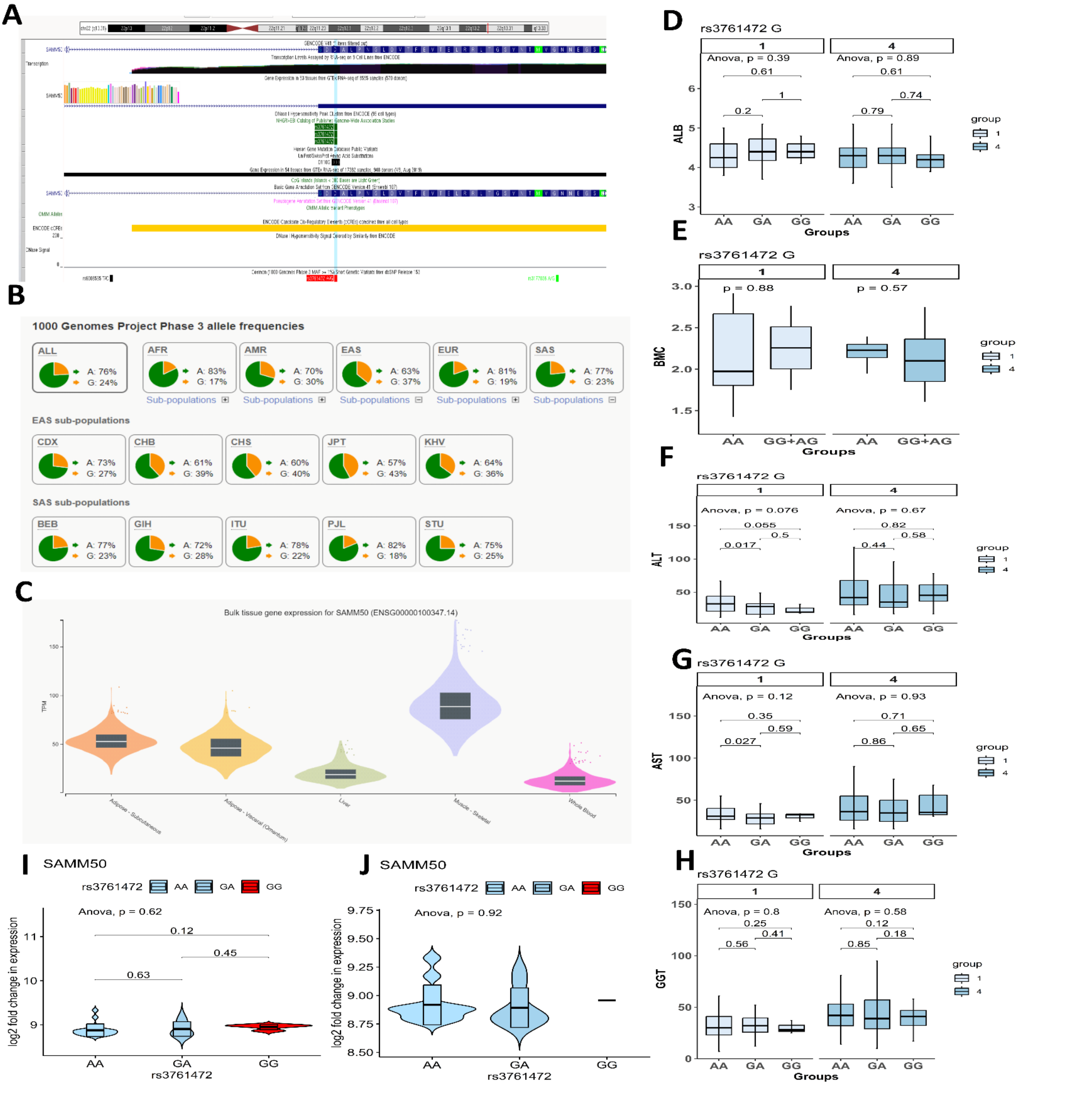

**Supplementary figure 3: Details of rs3761472**

A) Genomic annotation of rs3761472 in SAMM50 gene from dbSNP Short Genetic Variations (hg19) ; B) 1000 Genomes Project Phase 3 allele frequencies from ensembl.org ; C) Gene expression from GTEx portal for five different tissue types – adipose, skeletal and liver ; D-H) Distribution of liver damage markers and bone mineral content among different genotypes ;I- J) violin plots showing the effects of eQTL on SAMM50 gene expression in our study cohort.

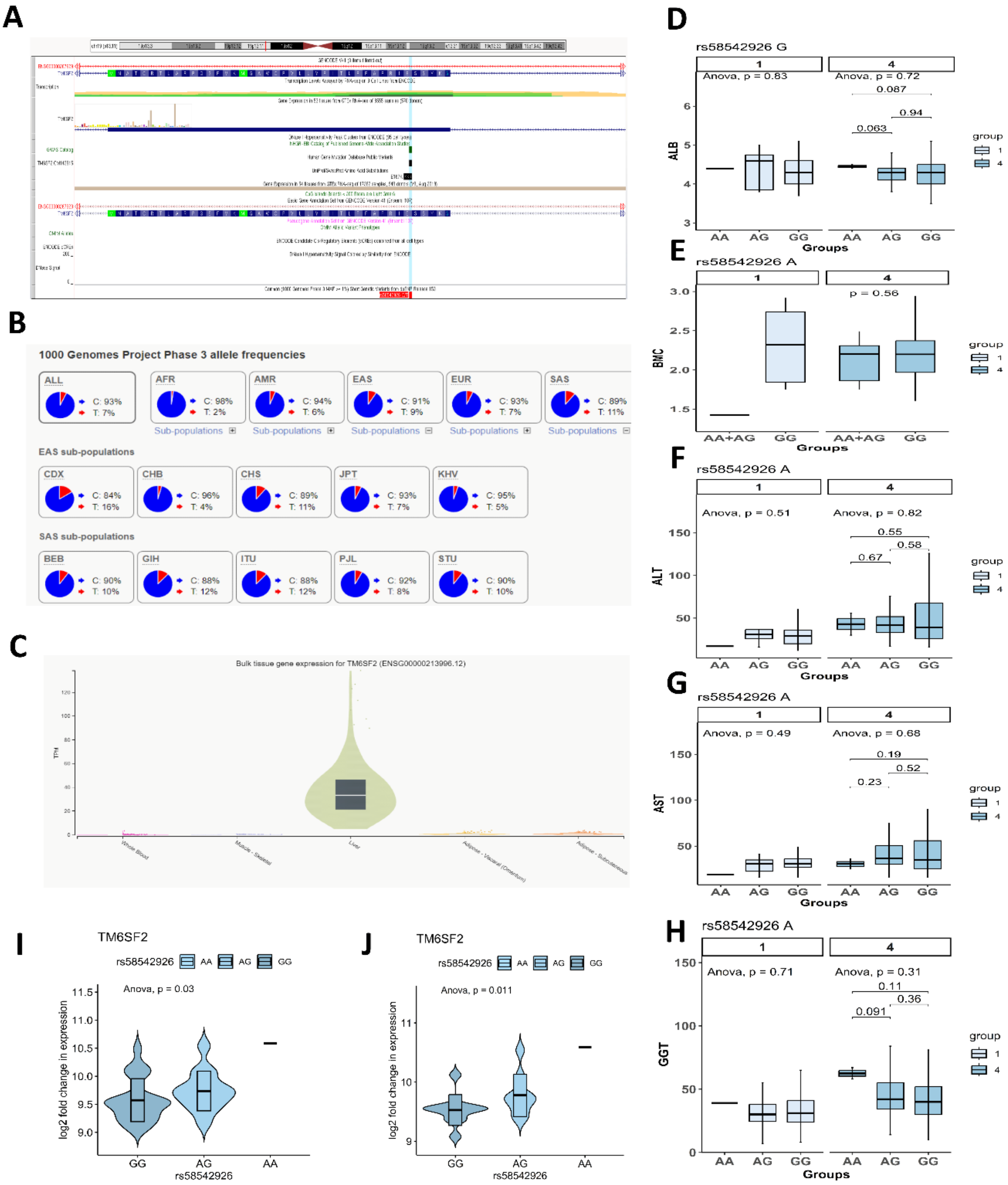

**Supplementary figure 4: Details of rs58542926**

A) Genomic annotation of rs58542926 in TM6SF2 gene from dbSNP Short Genetic Variations (hg19) ; B) 1000 Genomes Project Phase 3 allele frequencies from ensembl.org ;C) Gene expression from GTEx portal for five different tissue types – adipose, skeletal and liver; D-H) Distribution of liver damage markers and bone mineral content among different genotypes ;I- J) violin plots showing the effects of eQTL on TM6SF2 gene expression in our study cohort.

| Supplementary Table 1: Comparison of clinicopathological parametrs between fatty liver individuals and without fatty liver individuals | | | | |
|---|---|---|---|---|
| | **Parameters (unit)** | **FAT+ (n=260)** | **FAT- (n=291)** | **p-value** |
| | **Age (Years)** | 41.46±10.45 | 42.11±12.36 | 5.80E-01 |
| | **MALE:FEMALE** | 125:135 | 163:128 | |
| Anthropometry | **Weight(Kg)** | 66.87±9.95 | 58.2±11.48 | 7.29E-20 |
| | **BMI (Kg/m2)** | 26.97±3.85 | 23.13±4.3 | 5.95E-26 |
| | **HC (cm)** | 100.34±8.53 | 94.71±7.76 | 4.49E-12 |
| | **Waist circumference(cm)** | 93.2±8.6 | 84.13±11.3 | 1.66E-24 |
| Diabetes | **FBG (mg/dL)** | 96.08±36.13 | 85.81±19.83 | 0.00E+00 |
| | **Hba1c (mg/dL)** | 6.19±1.52 | 5.54±0.86 | 0.00E+00 |
| | **Fasting Insulin (IU/mL)** | 9.05±6.5 | 6.98±6.21 | 1.00E-03 |
| | **HOMAIR** | 2.24±2.48 | 1.58±2.46 | 7.00E-03 |
| Liver damage marker | **ALT (IU/l)** | 54.25±38.83" | 34.05±21.07 | 0.00E+00 |
| | **AST (IU/l)** | 43.58±26.69 | 31.24±11.51 | 0.00E+00 |
| | **GGT (IU/l)** | 46.6±29.08 | 37.09±21.67 | 2.24E-05 |
| Lipid level | **Triglyceride (mg/dL)** | 165.11±96.05 | 144.54±88.73 | 9.87E-03 |
| | **Total cholesterol (mg/dL)** | 183.33±43.58 | 169.74±41.36 | 2.20E-04 |
| | **HDL (mg/dL)** | 39.13±9.2 | 42.43±11.63 | 2.40E-04 |
| | **LDL (mg/dL)** | 111.73±41.45 | 97.77±38.59 | 6.25E-05 |
| | **VLDL (mg/dL)** | 31.47±13.08 | 27.68±11.67 | 4.53E-04 |
| | **% with metabolic dysregulation** | 80.99 | 57.88 | |
| Non-invasisve Fibrosis marker | **LSM (kPa)** | 6.45±3.2 | 5.25±3.63 | 1.81E-04 |
| Inflammation marker | **CRP (mg/dL)** | 4.9±3.81 | 3.7±3.29 | 8.95E-04 |
| Blood pressure | **SBP (mmHg)** | 126.11±17.13 | 124.7±18.67 | 3.60E-01 |
| | **DBP (mmHg)** | 80.36±11.16 | 77.97±11.52 | 1.53E-02 |
| | **Urea (mh/dL** | 18.52 ± 6.22 | 18.65 ± 6.23 | 2.40E-01 |
| | **Creatinine (mg/dL)** | 0.97±0.17 | 0.98±0.19 | 6.10E-01 |
| | **SS(mm)** | 2.95±1.82 | 2.11±2.06 | 5.39E-07 |
| | **BSP(mm)** | 1.29±1.06 | 0.88±0.64 | **1.17E-07** |
| | **TSP(mm)** | 1.9±1.45 | 1.36±1.08 | 1.88E-06 |
| | **NU(mm)** | 2.14±1.37 | 1.69±1.22 | 7.71E-05 |
| | **SFT(mm)** | 8.24±5.37 | 6.05±4.51 | 3.71E-07 |

**Suppl. Table 1:**

Comparison of clinicopathological parameters between fatty liver individuals and without fatty liver individuals

| Supplementry Table 2: Characteristics of body fat and muscle composition among the study groups | | | | | | | | | | | | |
|---|---|---|---|---|---|---|---|---|---|---|---|---|
| Total (n=112) | | | | | | | Non-obese | | | Obese | | |
| parameters (Units) | Group 4 (n=60) | Group 3 (n=14) | Group 2 (n=31) | Group 1 (n=7) | p-value | p-value (adjusted for covariates) | Group 4 | Group 3 | p-value | Group 4 | Group 3 | p-value |
| ***TBF.DEXA.PC*** | 49.06±18.47 | 39.84±14.83 | 41.05±14.28 | 38.41+11.91 | **0.055** | 1.044e-06 | 32.23±8.16 | 29.74±3.71 | 0.368 | 54.67±17.53 | 45.46±15.84 | 0.143 |
| ***Android.fat*** | 58.59±17.12 | 50.9±13.71 | 51.38±14.17 | 49.14+12.96 | **0.091** | 3.69E-05 | 44.49±8.38 | 42.18±5.49 | 0.494 | 63.4±16.72 | 55.74±14.71 | 0.189 |
| ***Gynoid.fat*** | 52.54±20.11 | 42.27±13.85 | 43.78±15.6 | 43.94+11.21 | **0.068** | 1.19E-06 | 34.47±8.45 | 33.68±3.51 | 0.773 | 58.7±19.22 | 47.04±15.28 | 0.067 |
| ***TISSUE.MASS*** | 66.6±10.89 | 65.79±9.04 | 62.13±8.38 | 54.3+9.8 | **0.0** | 1.973e-10 | 56.82±7.52 | 58.49±5.46 | 0.605 | 69.94±9.84 | 69.84±8.13 | 0.976 |
| ***FAT.MASS*** | 33.74±16.32 | 26.74±12.22 | 26.44±11.43 | 20.48+5.63 | **0.021** | 1.2e-06 | 18.39±5.26 | 17.38±2.69 | 0.587 | 38.85±15.53 | 31.93±12.4 | 0.167 |
| ***LEAN.MASS*** | 32.9±11.34 | 39.05±9.57 | 36.24±9.02 | 33.82+10.33 | 0.190 | 5.34E-04 | 38.43±6.26 | 41.11±4.67 | 0.336 | 31.01±12.1 | 37.91±11.57 | 0.132 |
| ***FAT.FREE.MASS*** | 34.87±11.95 | 41.52±10.12 | 38.4±9.48 | 35.98+10.84 | 0.176 | 5.83E-04 | 40.72±6.53 | 43.77±4.87 | 0.296 | 32.87±12.76 | 40.27±12.24 | 0.127 |
| ***BMC*** | 1.97±0.68 | 2.47±0.66 | 2.16±0.52 | 2.19+0.59 | **0.055** | 0.006208 | 2.29±0.34 | 2.67±0.34 | 0.074 | 1.86±0.73 | 2.36±0.78 | 0.101 |

**Suppl. Table 2:**

Characteristics of body fat and muscle composition among the study groups

| Supplementary table 3: Details of the associated SNPs | | | | | | | | | | | | | | | | | | |
|---|---|---|---|---|---|---|---|---|---|---|---|---|---|---|---|---|---|---|
| | | | | | | | | | | Risk allele frequency (MAF) | | | | 1000 Genome | | | | |
| SNP | Gene | CHR | BP | Alleles | Risk allele | Type of nucleotide change | p-value | OR | RsID | Group 4 | Group 3 | Group 2 | Group 1 | African | East Asian | Europe | South Asian | American |
| rs471364 | TTC39B | 9 | 15289578 | [T/C] | G | Silent | 0.001 | 3.076 [2.0 | rs471364 | 0.284 | 0.21 | 0.205 | 0.358 | C=0.2179 | C=0.0198 | C=0.1044 | C=0.044 | C=0.102 |
| rs2281135 | PNPLA3 | 22 | 44332570 | [A/G] | A | Silent | 0.001 | 2.911[2.02 | rs2281135 | 0.266 | 0.355 | 0.393 | 0.25 | A=0.1392 | A=0.3641 | A=0.1988 | A=0.239 | A=0.435 |
| rs7200543 | PDXDC1 | 16 | 15129970 | [A/G] | A | Synonymous_L7 | 0.031 | 2.26[1.08 | rs7200543 | 0.416 | 0.387 | 0.442 | 0.473 | G=0.1846 | G=0.3710 | G=0.3062 | G=0.442 | G=0.533 |
| rs3761472 | SAMM50 | 22 | 44368122 | [A/G] | G | Missense_D110 | 0.002 | 2.863 [2.0 | rs3761472 | 0.302 | 0.484 | 0.478 | 0.176 | G=0.1710 | G=0.3710 | G=0.1879 | G=0.233 | G=0.297 |
| rs10067427 | | 5 | 99342047 | [A/G] | G | | 0.003 | 0.8541 [0. | rs1006742 | 0.159 | 0.032 | 0.013 | 0.196 | A=0.2950 | A=0.7440 | A=0.6173 | A=0.810 | A=0.674 |
| rs738409 | PNPLA3 | 22 | 44324727 | [C/G] | G | Missense_I148 | 0.003 | 2.85[1.985 | rs738409 | 0.317 | 0.419 | 0.429 | 0.176 | G=0.1180 | G=0.3502 | G=0.2256 | G=0.246 | G=0.484 |
| rs1020689 | ALMS1P | 2 | 73900900 | [T/C] | G | Silent | 0.003 | 2.759[1.92 | rs1020689 | 0.392 | 0.129 | 0.188 | 0.203 | C=0.6180 | C=0.0060 | C=0.2078 | C=0.139 | C=0.259 |
| rs2068888 | | 10 | 94839642 | [A/G] | G | | 0.005 | 0.869 [0.8 | rs2068888 | 0.332 | 0.065 | 0.063 | 0.426 | G=0.7428 | G=0.2083 | G=0.5139 | G=0.310 | G=0.533 |
| rs6591182 | EHBP1L1 | 11 | 65349756 | [T/G] | A | Missense_V538 | 0.010 | 1.035 [1.0 | rs6591182 | 0.494 | 0.21 | 0.161 | 0.432 | G=0.3101 | G=0.4514 | G=0.5060 | G=0.508 | G=0.378 |
| rs1532085 | | 15 | 58683366 | [A/G] | G | | 0.014 | 1.034 [1.02 ± 1.047] | rs1532085 | 0.44 | 0.419 | 0.388 | 0.412 | A=0.5212 | A=0.4940 | A=0.3648 | A=0.559 | A=0.379 |
| rs5854292 | TM6SF2 | 19 | 19379549 | [T/C] | A | Missense_E167 | 0.021 | 2.735 [1.8 | rs5854292 | 0.491 | 0.032 | 0.116 | 0.5 | T=0.0227 | T=0.0863 | T=0.0676 | T=0.107 | T=0.063 |
| rs1088935 | DOCK7 | 1 | 63118196 | [A/C] | C | Silent | 0.026 | 2.763 [1.9 | rs1088935 | 0.416 | 0.339 | 0.406 | 0.324 | C=0.3918 | C=0.1766 | C=0.3012 | C=0.429 | C=0.344 |
| rs1832007 | AKR1C4 | 10 | 5254847 | [A/G] | G | Silent | 0.027 | 0.8627 [0. | rs1832007 | 0.105 | 0.032 | 0.022 | 0.074 | G=0.0250 | G=0.1111 | G=0.1203 | G=0.109 | G=0.202 |
| rs2800 | SLC9A9 | 3 | 1.43E+08 | [T/C] | G | Silent | 0.028 | 3.007 [2.0 | rs2800 | 0.437 | 0.016 | 0.018 | 0.432 | C=0.2950 | C=0.5972 | C=0.3400 | C=0.414 | C=0.357 |
| rs571312 | | 18 | 57839769 | [A/C] | A | | 0.036 | 1.052 [1.0 | rs571312 | 0.323 | 0.032 | 0.152 | 0.243 | A=0.3691 | A=0.1815 | A=0.2396 | A=0.318 | A=0.128 |
| rs3566508 | CECR5 | 22 | 17625915 | [A/G] | A | Missense_T149 | 0.038 | 2.783 [1.9 | rs3566508 | 0.012 | 0.387 | 0.433 | 0.047 | A=0.0015 | A=0.0000 | A=0.0477 | A=0.020 | A=0.029 |
| rs7805747 | PRKAG2 | 7 | 1.51E+08 | [A/G] | A | Silent | 0.040 | 2.758 [1.9 | rs7805747 | 0.066 | 0.452 | 0.451 | 0.041 | A=0.3245 | A=0.0030 | A=0.2883 | A=0.071 | A=0.173 |
| rs972283 | | 7 | 1.30E+08 | [A/G] | A | | 0.041 | 1.052 [1.0 | rs972283 | 0.377 | 0 | 0.018 | 0.473 | A=0.1233 | A=0.3155 | A=0.4394 | A=0.372 | A=0.360 |
| rs1040196 | SUGP1 | 19 | 19407718 | [T/C] | G | Silent | 0.043 | 2.72 [1.89 | rs1040196 | 0.162 | 0.306 | 0.379 | 0.095 | C=0.1808 | C=0.1071 | C=0.0706 | C=0.117 | C=0.082 |
| rs7703051 | | 5 | 74625487 | [A/C] | C | | 0.044 | 1.053 [1.0 | rs7703051 | 0.41 | 0.258 | 0.299 | 0.345 | C=0.1808 | C=0.1071 | C=0.0706 | C=0.117 | C=0.082 |
| rs7121446 | | 11 | 1.22E+08 | [A/G] | A | | 0.044 | 1.047 [1.0 | rs7121446 | 0.249 | 0.242 | 0.188 | 0.189 | A=0.3888 | A=0.0585 | A=0.2416 | A=0.248 | A=0.164 |

**Suppl. Table 3:**

Details of the associated SNPs.
